# Development of interoperable, domain-specific extensions for the German Corona Consensus (GECCO) COVID-19 research dataset using an interdisciplinary, consensus-based workflow

**DOI:** 10.1101/2022.05.12.22274089

**Authors:** Gregor Lichtner, Thomas Haese, Sally Brose, Larissa Röhrig, Liudmila Lysyakova, Stefanie Rudolph, Maria Uebe, Julian Sass, Alexander Bartschke, David Hillus, Florian Kurth, Leif Erik Sander, Falk Eckart, Nicole Toepfner, Reinhard Berner, Anna Frey, Marcus Dörr, Jörg Janne Vehreschild, Christof von Kalle, Sylvia Thun

**Affiliations:** Berlin Institute of Health at Charité – Universitätsmedizin Berlin, Berlin, Germany; Charité – Universitätsmedizin Berlin, Corporate Member of Freie Universität Berlin and Humboldt-Universität zu Berlin, Institute of Medical Informatics, Berlin, Germany; Universitätsmedizin Greifswald, Department of Anesthesia, Critical Care, Emergency and Pain Medicine, Greifswald, Germany; Robert Koch Institute, Department of Methodology and Research Infrastructure, Research Data Management, Berlin, Germany; National Association of Statutory Health Insurance Physicians (“Kassenärztliche Bundesvereinigung”; KBV), Digitalization and IT, Department Interoperability, Berlin, Germany; Charité - Universitätsmedizin Berlin, Corporate Member of Freie Universität Berlin and Humboldt-Universität zu Berlin, joint Charité and BIH Clinical Study Center, Berlin, Germany; Berlin Institute of Health at Charité – Universitätsmedizin Berlin, joint Charité and BIH Clinical Study Center, Berlin, Germany; Charité – Universitätsmedizin Berlin, Corporate Member of Freie Universität Berlin and Humboldt-Universität zu Berlin, Department of Infectious Diseases and Respiratory Medicine, Berlin, Germany; University Medical Centre Hamburg-Eppendorf, Department of Tropical Medicine, Bernhard Nocht Institute for Tropical Medicine and Department of Medicine I, Hamburg, Germany; Department of Pediatrics, University Hospital Carl Gustav Carus, Technische Universität Dresden, Dresden, Germany; University Hospital of Würzburg, Medical Clinic and Policlinic I, Würzburg, Germany; Universitätsmedizin Greifswald, Department of Internal Medicine B, Greifswald, Germany; Department I of Internal Medicine, University Hospital of Cologne, Cologne, Germany; German Centre for Infection Research (DZIF), Partner Site Bonn-Cologne, Cologne, Germany; Department II of Internal Medicine, Hematology/Oncology, Goethe University, Frankfurt, Frankfurt am Main, Germany

**Keywords:** COVID-19, Interoperability, GECCO dataset, FHIR, Research dataset, FAIR principles

## Abstract

**Background:** The COVID-19 pandemic has spurred large-scale, inter-institutional research efforts. To enable these efforts, researchers must agree on dataset definitions that not only cover all elements relevant to the respective medical specialty but that are also syntactically and semantically interoperable. Following such an effort, the German Corona Consensus (GECCO) dataset has been developed previously as a harmonized, interoperable collection of the most relevant data elements for COVID-19-related patient research. As GECCO has been developed as a compact core dataset across all medical fields, the focused research within particular medical domains demands the definition of extension modules that include those data elements that are most relevant to the research performed in these individual medical specialties.

**Objective:** To (i) specify a workflow for the development of interoperable dataset definitions that involves a close collaboration between medical experts and information scientists and to (ii) apply the workflow to develop dataset definitions that include data elements most relevant to COVID-19-related patient research in *immunization, pediatrics*, and *cardiology*.

**Methods:** We developed a workflow to create dataset definitions that are (i) content-wise as relevant as possible to a specific field of study and (ii) universally usable across computer systems, institutions, and countries, i.e., interoperable. We then gathered medical experts from three specialties (*immunization, pediatrics*, and *cardiology*) to the select data elements most relevant to COVID-19-related patient research in the respective specialty. We mapped the data elements to international standardized vocabularies and created data exchange specifications using HL7 FHIR. All steps were performed in close interdisciplinary collaboration between medical domain experts and medical information scientists. The profiles and vocabulary mappings were syntactically and semantically validated in a two-stage process.

**Results:** We created GECCO extension modules for the *immunization, pediatrics*, and *cardiology* domains with respect to the pandemic requests. The data elements included in each of these modules were selected according to the here developed consensus-based workflow by medical experts from the respective specialty to ensure that the contents are aligned with the respective research needs. We defined dataset specifications for a total number of 48 (*immunization*), 150 (*pediatrics*), and 52 (*cardiology*) data elements that complement the GECCO core dataset. We created and published implementation guides and example implementations as well as dataset annotations for each extension module.

**Conclusions:** These here presented GECCO extension modules, which contain data elements most relevant to COVID-19-related patient research in *immunization, pediatrics* and *cardiology*, were defined in an interdisciplinary, iterative, consensus-based workflow that may serve as a blueprint for the development of further dataset definitions. The GECCO extension modules provide a standardized and harmonized definition of specialty-related datasets that can help to enable inter-institutional and cross-country COVID-19 research in these specialties.

## Introduction

The COVID-19 pandemic has led to unprecedented strong efforts in connecting nationwide and international research to help in managing the disease and its effects on public health. To enable research across different health care providers, institutions or even countries, interoperability between the medical data systems is essential [1]. Therefore, early in the pandemic, the German Corona Consensus Dataset (GECCO) has been developed in a collaborative effort to provide a standardized, unified core dataset for inter-institutional COVID-19-related patient research [2]. The GECCO dataset specifies a set of 81 essential clinical data elements from 13 domains such as anamnesis & risk factors, symptoms, and vital signs, that have been selected by expert committees from university hospitals, professional associations, and research initiatives. Since its development, the GECCO dataset has been implemented in a large number of institutions, most notably in virtually every German university hospital, which now provides access to the GECCO dataset in the context of the German COVID-19 Research Network of University Medicine (“Netzwerk Universitätsmedizin”) [3,4].

The GECCO dataset has been developed to contain as many relevant data elements as possible, but few enough to keep the effort of implementing the dataset manageable. Therefore, the dataset contains mostly data elements of general research interest, excluding data elements that are only of interest for particular medical specialties or use cases. These data items are considered part of domain-specific extension modules to the GECCO dataset, which are introduced in this article.

Thus, we here aimed to develop domain-specific extensions to the GECCO dataset that cover the most relevant data elements for COVID-19-related patient research for the medical specialties of *immunization, pediatrics*, and *cardiology*. To that end, we first developed a workflow that aims at providing dataset definitions that (i) contain the most relevant data elements for the research aims of the end users and (ii) that can be applied universally across institutions and countries. We then followed that workflow with different groups of medical experts from different medical specialties to define extension modules relevant for *immunization, pediatrics*, and *cardiology*.

These extension modules complement the GECCO core dataset and use the same international health IT standards and terminologies as the GECCO dataset, such as the *Systematized Nomenclature of Medicine - Clinical Terms* (SNOMED CT)[5] and *Logical Observation Identifiers Names and Codes* (LOINC)[6,7] and the *Fast Healthcare Interoperability Resources* (FHIR)[8,9] standard. The extension modules were developed in close alignment with the GECCO dataset to ensure interoperability and compatibility with existing definitions.

We here describe the consensus-based data element selection and data format definition workflow that we applied in close collaboration between medical experts from *immunology, pediatrics*, and *cardiology* domains on the content definition side and medical information specialists and FHIR developers on the technical side. This workflow may serve as a blueprint for further development of consensus-based data set definitions.

## Methods

### Workflow definition

We aimed to develop a workflow to create dataset definitions that are (i) content-wise as relevant as possible to a specific field of study and (ii) universally usable across computer systems, institutions, and countries, i.e., interoperable. We based the specification of the workflow on our experience with the definition of the German Corona Consensus (GECCO) dataset, where health professionals from 50 institutions (university hospitals, professional associations and other relevant organizations) participated to define the most relevant data elements for general scope COVID-19-related research [2]. To fulfil the first requirement (relevancy), we decided to leave the full responsibility of data element selection to groups of medical professionals of the respective specialty, with only minimal interference by the development team. We have deliberately left the exact process open of how the group of medical experts may select the data elements (e.g., literature review, focus groups, consensus-based processes) to allow maximal flexibility of the dataset definition workflow with respect to the medical experts’ values and preferences. To fulfil the second requirement (interoperability), we adopted a model loosely based on Jacobsen’s workflow for data FAIRification [10], with mapping, quality assurance and publication steps as outlined in detail below.

### Selection of data items

The content of the domain-specific research datasets was defined by medical domain experts in a transparent workflow (Figure 1). The involvement of the medical domain experts as the end-users of the data to be provided ensured that the contents of the datasets are aligned to the actual research needs. In our project, the so-called subject- and organ-specific working groups of the national pandemic cohort net (NAPKON) served as the domain-specific groups of medical experts. These groups were established by voluntary association of medical experts from the respective medical specialty in the context of the nationwide NAPKON project in Germany. Each of the subject- and organ-specific working groups elected a board, and all communication between the dataset developers and the working groups was organized and carried out via the working groups’ board. In preparation for the GECCO extension modules, we invited the subject- and organ-specific groups for *immunology, pediatrics* and *cardiology* to provide up to 50 data elements with up to 10 response items each that were, in the view of the medical experts, the most relevant data elements to patient-related COVID-19 research in their medical specialty and that were not already included in the GECCO core dataset.

**Figure 1.**
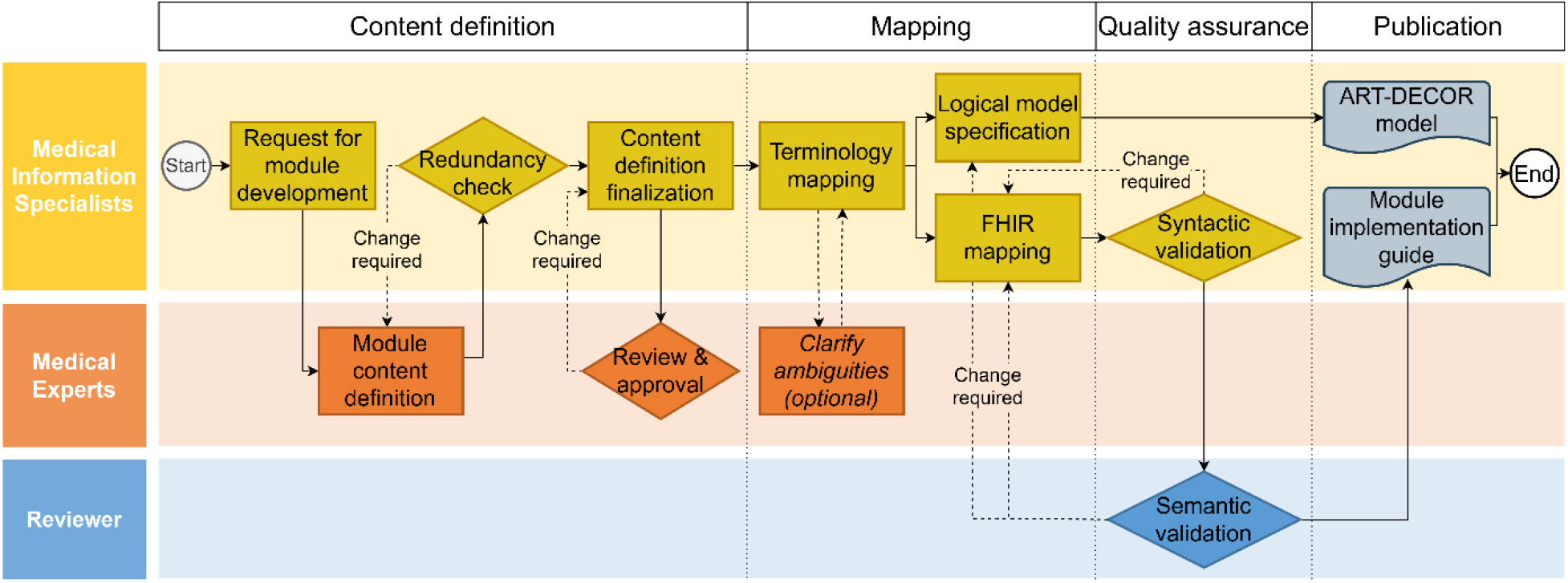
Flowchart of the consensus-based, interdisciplinary dataset definition and mapping workflow for the domain-specific COVID-19 research datasets.

If necessary, more data items or response options could be provided in coordination with the development team. The provided data items were then reviewed by the development team and a first definition of the contents of the extension module was returned to the respective subject- and organ-specific working group for approval or change requests. After approval by the subject- and organ-specific working group, the definition of the extension module content was considered finalized.

### Development of the standardized data formats

To map the data items selected by the subject- and organ-specific working groups to international standard vocabularies, we performed a consensus-based mapping procedure, where every concept was mapped to appropriate vocabularies SNOMED CT for general concepts [11], LOINC for observations [7], *International Statistical Classification of Diseases and Related Health Problems, 10*^*th*^ *revision, German modification* (ICD-10-GM) for diagnoses [12], *Anatomical Therapeutic Chemical Classification System* (ATC) for Germany for drugs and active ingredients [13], *Unified Code for Units of Measure* (UCUM) for measurement units [14]) by two medical information scientists independently. Ambiguities and non-matching mappings were then discussed within the development team and in close collaboration with the medical experts of the subject- and organ-specific working groups until consensus was achieved. The data item-to-concept mappings were annotated on ART-DECOR, an open-source collaboration platform for creating and maintaining dataset element descriptions [15].

As for the GECCO dataset, the format for data exchange was specified using HL7 FHIR resources. The mapping of data items to FHIR resources was performed in an iterative, consensus-based workflow among the development team. Wherever possible, published FHIR profiles from the GECCO dataset, from the Medical Informatics Initiative (MII) [16] or the National Association of Statutory Health Insurance Physicians (“Kassenärztliche Bundesvereinigung”; KBV) [17] – in this order of priority – served as the base definition for the future extension module profiles.

The profiles and value sets were specified using the FHIR Shorthand (FSH) language (version 1.2.0) and translated to Structure Definition JSON files using the HL7 FSH SUSHI software package (version 2.2.3) [18,19]. We required that at least one exemplary instance be defined for every profile. Syntactic validation of the profiles and value sets definitions was performed using the error-free conversion of the FSH files to JSON using SUSHI and subsequent validation of each profile with their defined instances using the HL7 FHIR validator as implemented in the FHIR Shorthand Validator Python package (version 0.2.2) [20]. After successful syntactic validation of a set of profiles, the profiles were subjected to a two-stage review process as follows. First, the profiles and corresponding value sets and extensions were internally reviewed for semantic appropriateness with the GECCO core developer (JS). After all necessary changes and approval by the internal reviewer, the profiles were subjected to the second review round by an external FHIR development expert. Subsequent to necessary corrections and approval of the external reviewer, the respective profiles together with their value sets and optionally extensions and code systems were considered finalized and published to the main branch of the git repository.

The whole development process was performed collaboratively on GitHub. Syntactic validation of the profiles was performed by continuous integration/continuous development (CI/CD) workflows implemented as GitHub actions. Semantic validation during the internal and external review rounds was performed using pull requests to two different git branches. After the final approval, profiles and value sets were merged into the main branch of the extension module’s repository, which served as the publication branch of the respective module. Since then, maintenance requests and updates of the extension modules are handled via GitHub issues. All kinds of relevant changes become a subject of the internal review as defined above; major changes (e.g., non-technical corrections) are additionally exposed to the external review.

Implementation guides were created for all three extension modules using the FHIR IG publisher tool and a customized template for the implementation guide’s HTML pages [21]. The implementation guides are published to GitHub pages and remain automatically synchronized with the main branch of the respective repository via CI/CD workflows.

## Results

### Dataset definition workflow

We developed an interdisciplinary, iterative, consensus-based workflow for the definition of domain-specific COVID-19 research datasets based on two key requirements: The first key requirement for the content of the datasets was that the content definition (i.e., selection of data elements) was to be performed in full responsibility by a group of medical experts to ensure that the selected data elements are truly those that are required in research of their respective medical specialty. The second key requirement was to produce FAIR (Findable, Accessible, Interoperable, Reusable) digital assets [22]: The dataset definitions shall be represented in FHIR profiles and implementation guides and these shall be registered on open platforms (Findable), they shall be retrievable through open, free, standard protocols (Accessible), they shall use only standard, international medical terminologies such as SNOMED CT and LOINC (Interoperable) and they shall be released with rich usage guides and examples (FHIR implementation guide) and under a permissive license (Reusable).

To fulfill these requirements, the dataset definition workflow consists of four phases: Content definition, mapping, quality assurance and publication (Figure 1). In the content definition phase, a group of medical experts from a particular medical specialty are approached by the development team consisting of medical information specialists and asked to provide a list of the data elements that are most relevant to patient-related COVID-19 research in their respective medical specialty. How the medical expert group compiles the list in detail is left to their discretion (e.g., based on systematic literature review, or Delphi consensus processes). The medical information scientists only review the provided lists for consistency and redundancy and compile the final content definition in agreement with the medical experts group. In the mapping phase, all data elements are then mapped to international terminologies in consultation with the group of medical experts. Based on these a logical model and the mappings of data elements to FHIR resources are established. In the quality assurance phase, the FHIR specifications are syntactically validated using automated software tools and then subjected to a two-staged review process with two individual data interoperability and harmonization experts to validate the specifications semantically, i.e., validate that the data elements defined by the group of medical experts are appropriately mapped to international standards. After any required changes, the logical model and the FHIR implementation guide are published openly accessible to the research community in repositories that fulfill the FAIR criteria as closely as possible, such as ART-DECOR[15] for logical models and GitHub or the FHIR Implementation Guide registry for the implementation guide[23].

### Datasets contents

#### Groups of medical experts

In the context of the national pandemic cohort net (“Nationales Pandemie Kohorten Netz”; NAPKON) project of the German COVID-19 Research Network of University Medicine [24], so-called subject- and organ-specific working groups were established by the voluntary association of medical experts from different medical specialties. In preparation for the domain-specific dataset definitions that extend the GECCO core dataset, the working groups for *immunology, pediatrics*, and *cardiology* were invited by the dataset development group to provide up to 50 data elements with up to 10 response items each that were of particular interest to their field concerning patient-related COVID-19 research and that were not already included in the GECCO core dataset. For the *immunization* dataset definition, physicians from the “NUM-COVIM” study for the determination and use of SARS-CoV-2 immunity [25–27] assumed the role of the organ-specific working group, as no such working group had been established previously.

#### Overview

The domain-specific dataset definitions developed in this work extend the GECCO core dataset by a total number of 48 data items for the *immunization* extension module, 150 for the *pediatrics* extension module, and 52 for the *cardiology* extension module. These data items have been collected via an iterative consensus-based approach from the respective subject- and organ-specific working groups and belonging to 10 of the 13 data categories of the GECCO dataset (Table 1). Data elements and number of items for each individual extension module are shown in Table 2, Table 3, and Table 4. The full lists of items are shown in the supplementary tables 1, 2, and 3.

**Table 1.** Number of data items per GECCO dataset category for each extension module.

| GECCO Data Category | GECCO Extension Module |  |  |
| --- | --- | --- | --- |
|  | Immunization | Pediatrics | Cardiology |
| Anamnesis & Risk factors | 13 | 21 | 6 |
| Complications | 24 | 47 | 7 |
| Demographics | - | 6 | - |
| Epidemiological factors | - | - | - |
| Imaging | - | 2 | 36 |
| Laboratory values | 1 | 27 | 2 |
| Medication | 1 | 35 | 1 |
| Onset of illness & admission | 6 | 2 | - |
| Outcome at discharge | - | - | - |
| Study enrollment & Inclusion criteria | - | - | - |
| Symptoms | - | 9 | - |
| Therapy | 2 | 1 | - |
| Vital signs | 1 | - | - |
| **Total items** | **48** | **150** | **52** |

**Table 2.** Types of data elements in the immunization extension module extending the GECCO core dataset. Shown are the data elements and the FHIR resource they have been mapped to, as well as the number of items for each data element (i.e., different response options).

| Category | Data Element | FHIR Resource | # items |
| --- | --- | --- | --- |
| Anamnesis | Chemotherapy | Procedure | 1 |
|  | Immunosuppressive therapy | Procedure | 1 |
|  | Regular Alcohol Intake | Observation | 2 |
| COVID-19 infection & treatment | Disease course | Encounter, Procedure | 5 |
|  | SARS-CoV-2 infection | Condition | 1 |
|  | SARS-CoV-2 variant | Observation | 1 |
| Immunization | Contraindications to immunization | Immunization | 2 |
|  | Immunizations performed | Immunization | 3 |
|  | Reason for immunization | Immunization | 5 |
|  | Willingness to receive additional immunization doses | Observation | 1 |
| Immunization reactions | Analgesic or antipyretic drug intake | MedicationStatement | 1 |
|  | Body temperature | Observation | 1 |
|  | Complications after immunization | Observation | 5 |
|  | Medical treatment for adverse reactions | Encounter | 3 |
|  | Symptoms after Vaccination | Condition | 16 |
| Total |  |  | 48 |

**Table 3.** Types of data elements in the pediatrics extension module extending the GECCO core dataset. Shown are the data elements and the FHIR resource they have been mapped to, as well as the number of items for each data element (i.e., different response options).

| Category | Data Element | FHIR Resource | # items |
| --- | --- | --- | --- |
| Complications | Complications to COVID-19 | Condition | 47 |
| Demographics | Body measures | Observation | 6 |
| Imaging | Echocardiography | Procedure, Imaging Study | 1 |
|  | PET-CT | Procedure, Imaging Study | 1 |
| Immunization | Immunizations performed | Immunization | 2 |
| Laboratory values | Laboratory values | Observation | 27 |
| Medical history | Chronic Hematologic Diseases | Condition | 8 |
|  | Chronic Kidney Diseases | Condition | 2 |
|  | Congenital Disease | Condition | 1 |
|  | Gastrointestinal Diseases | Condition | 6 |
|  | Medical History Stem Cells Transplant | Condition | 2 |
| Medication | Medication | MedicationStatement, List | 35 |
| Symptoms | COVID-19 Symptoms | Condition | 9 |
| Therapy | Hospitalization | Observation | 2 |
|  | Thoracic Drainage | Procedure | 1 |
| Total |  |  | 150 |

**Table 4.** Types of data elements in the cardiology extension module extending the GECCO core dataset. Shown are the data elements and the FHIR resource they have been mapped to, as well as the number of items for each data element (i.e., different response options).

| Category | Data Element | FHIR Resource | # items |
| --- | --- | --- | --- |
| **Anamnesis** | Chronic cardiologic diseases | Condition | 6 |
| **COVID-19-related complications** | Cardiologic complications of COVID-19 | Condition | 7 |
| **Echocardiography** | Echocardiography findings | Observation | 20 |
|  | Echocardiography procedure | Procedure | 3 |
| **Electrocardiography** | Electrocardiography findings | Observation | 11 |
|  | Electrocardiography procedure | Procedure | 2 |
| **Laboratory Values** | Laboratory values | Observation | 2 |
| **Medication** | Angiotensin receptor antagonist | MedicationStatement | 1 |
| **Total** |  |  | **52** |

All data items were mapped to the appropriate FHIR resources Observation, Condition, Procedure, MedicationStatement, Encounter, Questionnaire, QuestionnaireResponse, Immunization, ImagingStudy, List, and Specimen, and 26, 14, and 18 profiles (25, 17, and 12 value sets) were created for the *immunization, pediatrics*, and *cardiology* extension module, respectively. The data items that were already part of the GECCO dataset and that were not removed during the data selection step were taken over from GECCO and referenced as such in the implementation guides.

The implementation guides for the three extension module have been published on GitHub pages [28–30]. The source FHIR ShortHand (FSH) files have been published on GitHub [31–33]. Logical models and dataset descriptions are hosted on ART-DECOR, an open collaboration platform for modelling dataset definitions and their descriptions and terminology bindings [34–36].

## Discussion

We here present an interdisciplinary, iterative, consensus-based workflow to the definition of research datasets, focusing on creating datasets with the most relevant data elements for a particular field of study and on creating universally usable datasets according to the FAIR principles [22]. We applied the workflow to develop three GECCO extension modules that contain data items relevant for COVID-19-related patient research in the *immunization, pediatrics*, and *cardiology* fields. These extension modules complement the GECCO core dataset for domain-specified research. The data items are represented in HL7 FHIR profiles and use international terminologies, to ensure a harmonized, standardized, and interoperable dataset definition for these medical domains. The provision of data according to the extension modules introduced in this article will enable cross-institutional and cross-country data collection and collaborative research with a particular focus in *immunization, pediatrics*, and *cardiology*.

We have specified and implemented an interdisciplinary, iterative, consensus-based workflow for the selection of data items and the development of the dataset definition. The close collaboration and the constant feedback loops with domain experts from the respective medical specialties right from the beginning of the project, as performed here, are key for the successful development of a useful dataset definition. Indeed, since the selection of relevant data items was driven by the end-users of the dataset, who are the researchers that later will be using the data for their specialized areas of research, the semantic usability of the datasets is guaranteed. Likewise, having medical information specialists develop the formal dataset specification ensures technical interoperability and usability of the dataset definition.

Next to the successful development of dataset definitions, several factors determine a successful deployment or use of the developed extension modules [37]. First and most importantly clear and concise documentation of how to implement and provide data using the dataset definition is required. For FHIR-based dataset definitions, so-called implementation guides are used to provide both a narrative overview as well as technical details on the dataset definition [38]. Thus, we have created and published implementation guides for each of the here-developed extension modules. Second, the example implementations of the extension modules serve as a blueprint for developers and data engineers who implement the extension modules for their clinical databases. From our experience with the implementation of the GECCO dataset, well-defined example data items may be of equal if not higher importance than the technical description of the dataset specification, as developers and engineers tend to use the examples as blueprints for their implementation. Thus, we equipped every FHIR profile defined in the extension modules with at least one example. These examples are incorporated and issued within the implementation guides of the modules. Specifically, we aimed to provide one example for each different category of response option per profile. Thirdy, the actual implementation of the extension modules should be part of follow-up infrastructure projects to supply funding and resources for filling the dataset definition with actual data. For the GECCO dataset, this is ensured by follow-up projects of the German COVID-19 Research Network of University Medicine (“Netzwerk Universitätsmedizin”), such as CODEX+, which includes several implementation tasks that are actively using the GECCO dataset items [39] and further projects [40–43]. Fourth, once the dataset definitions are implemented and leveraged in use cases, additional demands to the dataset are likely raised or issues with existing definitions are revealed. The maintenance of existing definitions (e.g., performing technical corrections or even evolving the definitions or adding new items) is, therefore, necessary and must be organized and funded. Last, successful use of the extension modules is also highly dependent on the degree of interoperability of the dataset definitions in the first place [1,44,45]. For example, the use of questionnaires to assess certain features is common in clinical research. However, depending on the exact wording of the question and the number and wording of response options, results from different studies might not be directly comparable although they assessed the same features, as the questions and response options differ between studies. In the presented extension modules, several items were at first specified in a questionnaire-like fashion and direct implementation of these as Questionnaire resources in FHIR would have limited the applicability of such data elements, especially when aiming to map these elements from an electronic health records (EHR) system. In these cases, we revised the data elements specification to use interoperable concepts rather than questions. Here, repeated consultation with and final approval of the group of medical experts was key to be able to convert questions into interoperable concepts that convey the same information as intended by the content definition of the group of medical experts. In general, we recommend not to use Questionnaire/QuestionnaireResponse FHIR profiles in cases where the information to be represented can be modeled using more general, interoperable concepts and FHIR resources.

## Conclusion

We here introduce the development workflow and the resulting dataset definitions for GECCO extension modules for the *immunization, pediatrics*, and *cardiology* domains. We have defined and implemented a workflow in which interdisciplinary teams of medical domain experts, medical information scientists and FHIR developers closely collaborate in an iterative, consensus-based fashion for the successful development of useful and interoperable dataset definitions. This workflow may serve as a blueprint for further dataset definition projects, such as further dataset definitions for extending the GECCO core dataset. The extension modules described in this work have been validated and published. Their implementation and active use are anticipated in the context of current nationwide COVID-19 research networks in Germany.

## Data Availability

All data produced are available online at https://github.com/BIH-CEI

## Acknowledgements

The NAPKON (“Nationales Pandemie Kohorten Netz”, German National Pandemic Cohort Network) project is funded under a scheme issued by the Network of University Medicine (Nationales Forschungsnetzwerk der Universitätsmedizin (NUM)) by the Federal Ministry of Education and Research of Germany (Bundesministerium für Bildung und Forschung (BMBF)) grant number 01KX2021. The funding body did not take a role in the design of the study, development of the dataset or in the writing of the manuscript.

We thank Yannick Börner for his valuable contribution to the definition of the FHIR profiles. We thank all members of the subject- and organ-specific working groups.

## Data Availability

The implementation guides for the three extension modules have been published on GitHub pages [28–30]. The source FHIR ShortHand (FSH) files have been published on GitHub [31–33]. Dataset descriptions can be found on ART-DECOR [34–36]

## Authors’ contributions

All authors contributed to the development of the extension modules. GL, TH, SB, LR, JS, AB, ST performed terminology mapping, FHIR profiling and critical review of the concept and resource mappings. TH, SB, LR defined the datasets in ART-DECOR. DH, FK, LES, FB, FE, NT, RB, AF, MD developed and compiled the list of data items for the datasets. SR, LL and MU coordinated the project and the consensus finding process within and between working groups. JJV, CvK, ST conceived the work. GL drafted the manuscript. All authors read and approved the final manuscript.

## Supplementary Appendix

The following tables show the data items that are included in the immunization extension module (Table S 1), the pediatrics extension module (Table S 2), and the cardiology extension module (Table S 3). Note that these tables list only the data items of the extension modules that are not included in the GECCO core dataset and that the complete dataset definition for each specialty consists of the GECCO core dataset together with the data items of the extension module.

### GECCO Immunization extension module

**Table S 1.** Data items in the immunization extension module extending the GECCO core dataset. Shown are the data elements and the FHIR resource they have been mapped to, as well as the items for each data element (i.e., different response options).

| Category | Data Element | FHIR Resource | Item |
| --- | --- | --- | --- |
| **Anamnesis** | Chemotherapy | Procedure | Chemotherapy |
|  | Immunosuppressive therapy | Procedure | Immunosuppressive therapy |
|  | Regular Alcohol Intake | Observation | Frequency |
|  |  |  | Quantity |
| **COVID-19 infection & treatment** | Disease course | Encounter, Procedure | No symptoms |
|  |  |  | Treated at home |
|  |  |  | Treated at hospital with oxygen therapy |
|  |  |  | Treated at hospital without oxygen therapy |
|  |  |  | Treated at intensive care unit |
|  | SARS-CoV-2 infection | Condition | SARS-CoV-2 infection |
|  | SARS-CoV-2 variant | Observation | SARS-CoV-2 variant |
| **Immunization** | Contraindications to immunization | Immunization | Allergy |
|  |  |  | Pregnancy |
|  | Immunizations performed | Immunization | Date |
|  |  |  | Lot number |
|  |  |  | Type |
|  | Reason for immunization | Immunization | Everybody gets vaccinated |
|  |  |  | Protection of private environment from infection/disease |
|  |  |  | Protection of themself from infection/disease |
|  |  |  | Protection of work environment from infection/disease |
|  |  |  | Worrying about disadvantages |
|  | Willingness to receive additional immunization doses | Observation | Willingness to receive additional immunization doses |
| **Immunization reactions** | Analgesic or antipyretic drug intake | MedicationStatement | Analgesic or antipyretic drug intake |
|  | Body temperature | Observation | Body temperature after vaccination |
|  | Complications after immunization | Observation | Allergic reaction after immunization |
|  |  |  | Injection site erythema |
|  |  |  | Injection site pain at rest |
|  |  |  | Injection site pain during pressure/movement |
|  |  |  | Injection site swelling |
|  | Medical treatment for adverse reactions | Encounter | Ambulatory |
|  |  |  | Inpatient |
|  |  |  | No treatment |
|  | Symptoms after Vaccination | Condition | Chill |
|  |  |  | Diarrhea |
|  |  |  | Difficulty breathing |
|  |  |  | Dyspnea |
|  |  |  | Exhaustion |
|  |  |  | Fatigue |
|  |  |  | Feeling feverish |
|  |  |  | Fever |
|  |  |  | Fever with chills |
|  |  |  | Joint pain |
|  |  |  | Liquid stool |
|  |  |  | Loose stool |
|  |  |  | Muscle pain |
|  |  |  | Nausea |
|  |  |  | Soft stool |
|  |  |  | Vomiting symptom |

### GECCO Pediatrics extension module

**Table S 2.**
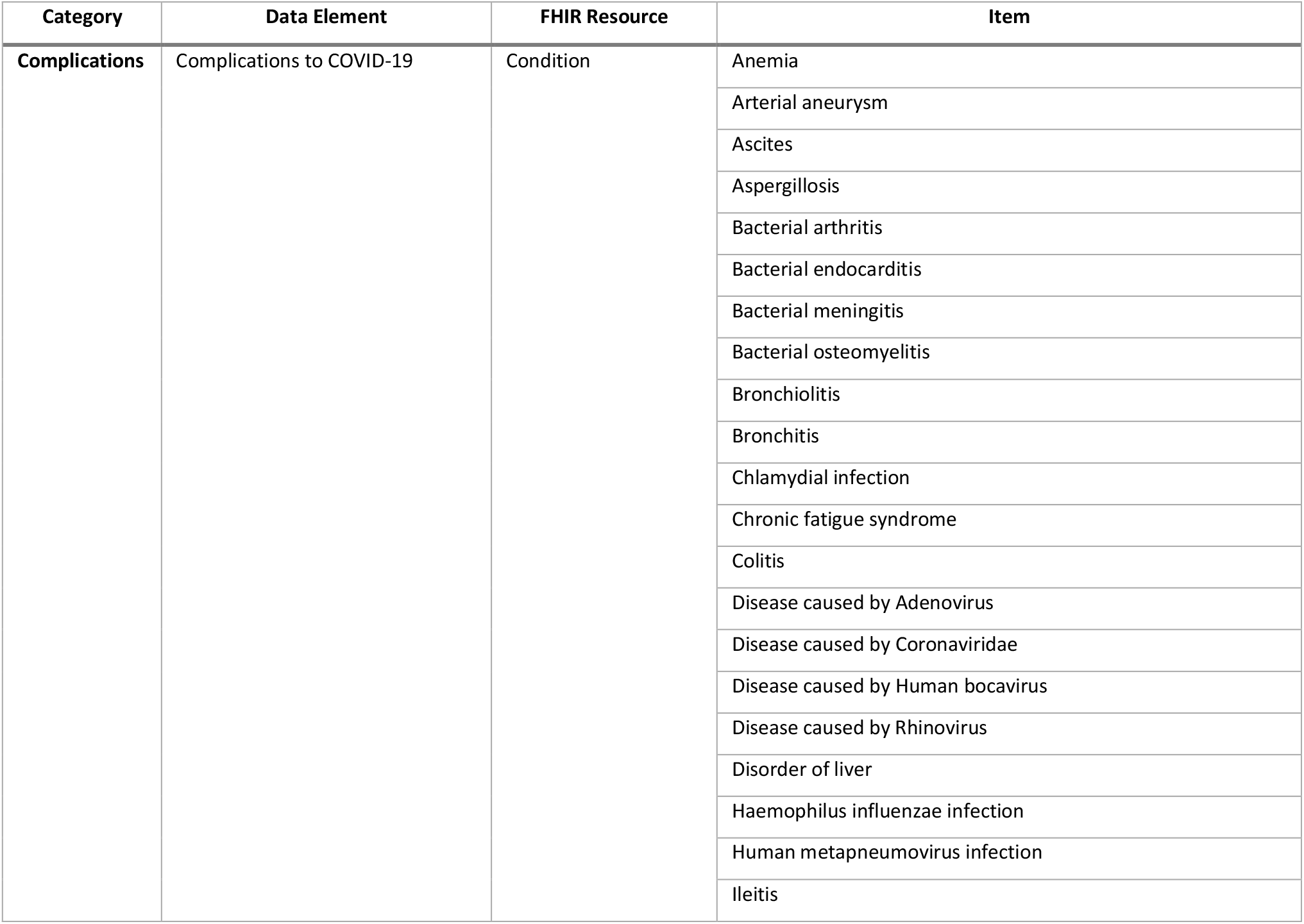
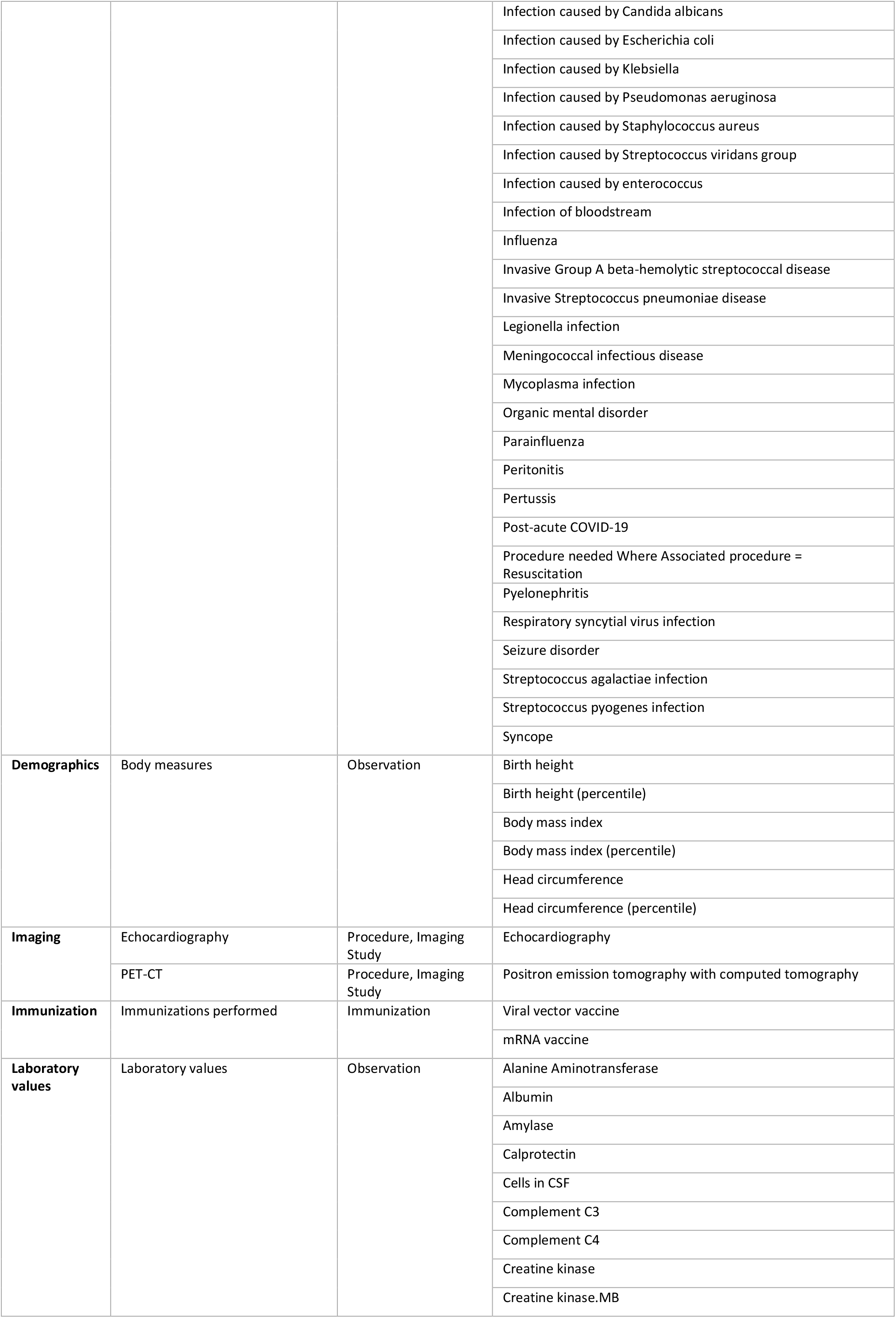
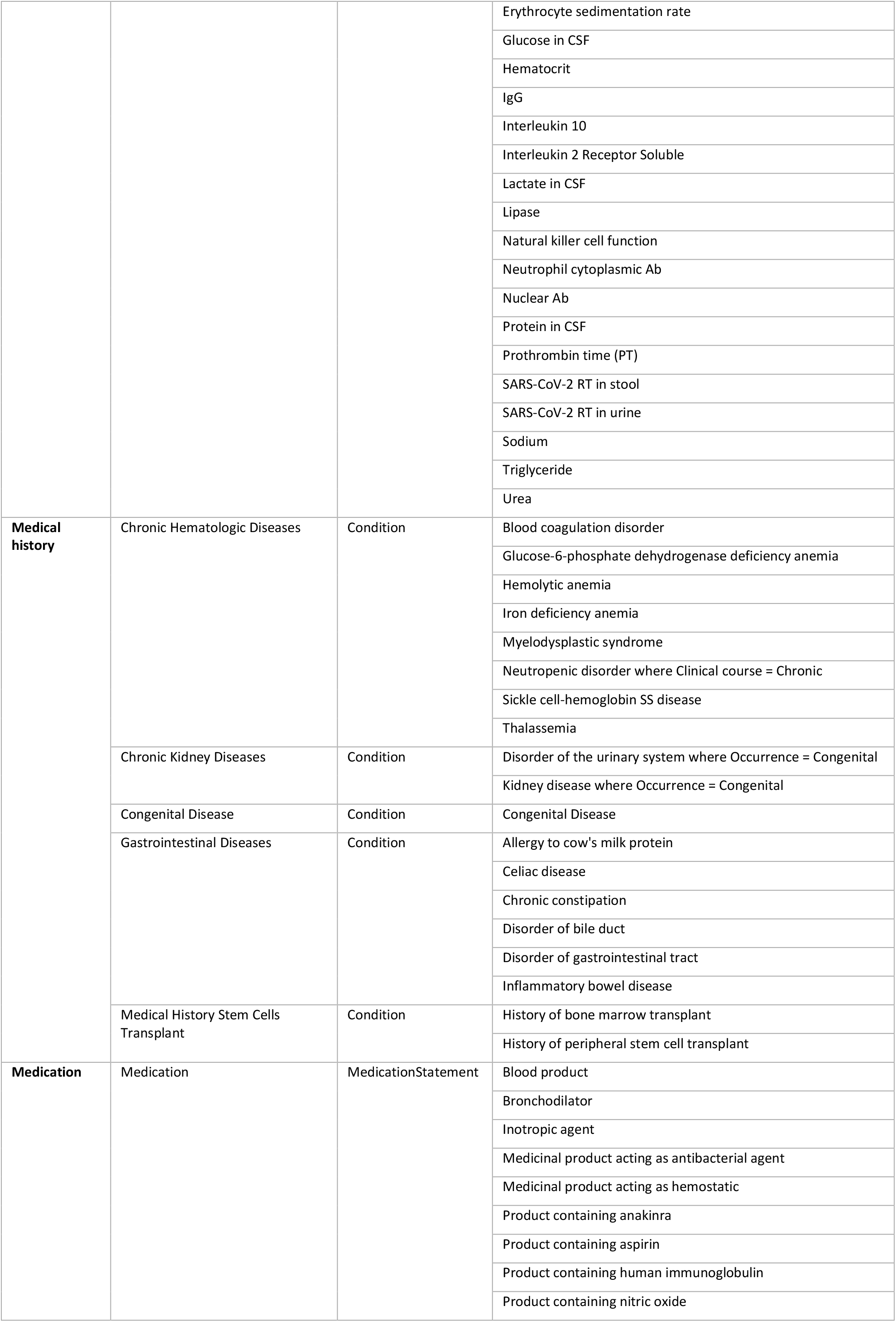
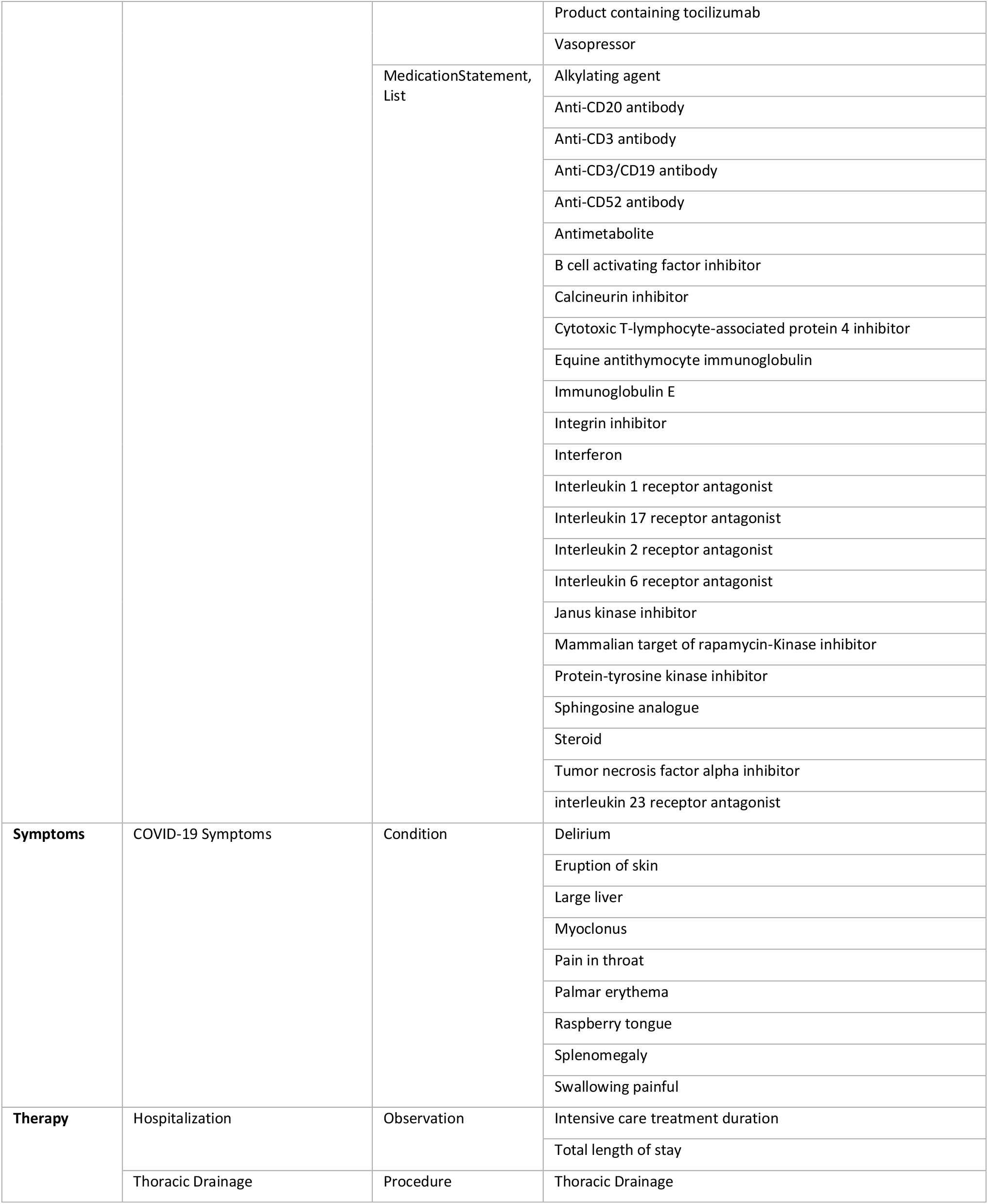
Data items in the pediatrics extension module extending the GECCO core dataset. Shown are the data elements and the FHIR resource they have been mapped to, as well as the items for each data element (i.e., different response options).

### GECCO Cardiology extension module

**Table S 3.** Data items in the cardiology extension module extending the GECCO core dataset. Shown are the data elements and the FHIR resource they have been mapped to, as well as the items for each data element (i.e., different response options).

| Category | Data Element | FHIR Resource | Item |
| --- | --- | --- | --- |
| Anamnesis | Chronic cardiologic diseases | Condition | Atrial fibrillation |
|  |  |  | Atrial flutter |
|  |  |  | Cardiomyopathy |
|  |  |  | Congenital heart disease |
|  |  |  | Heart failure |
|  |  |  | History of coronary artery bypass grafting |
| COVID-19-related complications | Cardiologic complications of COVID-19 | Condition | Bacterial respiratory infection |
|  |  |  | Cardiogenic shock |
|  |  |  | Complete atrioventricular block |
|  |  |  | Myocarditis |
|  |  |  | Pericardial effusion |
|  |  |  | Ventricular fibrillation |
|  |  |  | Viral disease |
| Echocardiography | Echocardiography findings | Observation | Abscess of heart |
|  |  |  | Aortic valve regurgitation |
|  |  |  | Aortic valve stenosis |
|  |  |  | Heart valve disorder |
|  |  |  | Left Ventricular Ejection Fraction |
|  |  |  | Left ventricular hypertrophy |
|  |  |  | Left ventricular wall motion abnormality |
|  |  |  | Mitral valve regurgitation |
|  |  |  | Mitral valve stenosis |
|  |  |  | Paradoxical cardiac wall motion |
|  |  |  | Pericardial effusion |
|  |  |  | Pulmonic valve regurgitation |
|  |  |  | Pulmonic valve stenosis |
|  |  |  | Right ventricular hypertrophy |
|  |  |  | Thrombosis |
|  |  |  | Tricuspid annular plane systolic excursion (TAPSE) |
|  |  |  | Tricuspid valve regurgitation |
|  |  |  | Tricuspid valve stenosis |
|  |  |  | Vegetation of heart |
|  |  |  | Ventricular hypertrophy |
|  | Echocardiography procedure | Procedure | Date |
|  |  |  | Echocardiography |
|  |  |  | Type of echocardiography |
| Electrocardiography | Electrocardiography findings | Observation | Atrial ectopics |
|  |  |  | Atrioventricular Block |
|  |  |  | Bundle Branch Block |
|  |  |  | Inverted T wave |
|  |  |  | Low QRS voltages |
|  |  |  | Premature ventricular contractions |
|  |  |  | QRS Axis |
|  |  |  | QRS Interval |
|  |  |  | QT Interval |
|  |  |  | ST Interval |
|  |  |  | Sinus rhythm |
|  | Electrocardiography procedure | Procedure | 12 lead electrocardiogram |
|  |  |  | Date |
| **Laboratory Values** | Laboratory values | Observation | Troponin I |
|  |  |  | Troponin T |
| **Medication** | Angiotensin receptor antagonist | MedicationStatement | Angiotensin Receptor Antagonist |

